# Long-term psychological outcomes following stroke: The OX-CHRONIC study

**DOI:** 10.1101/2023.03.27.23287789

**Authors:** Andrea Kusec, Elise Milosevich, Owen A. Williams, Evangeline G. Chiu, Pippa Watson, Chloe Carrick, Bogna A. Drozdowska, Avril Dillon, Trevor Jennings, Bloo Anderson, Helen Dawes, Shirley Thomas, Annapoorna Kuppuswamy, Sarah T. Pendlebury, Terence J. Quinn, Nele Demeyere

**Author notes:** Corresponding Author Address: Nuffield Department of Clinical Neurosciences, University of Oxford, John Radcliffe Hospital, Oxford, OX3 9DU, UK. Patient and Public Involvement Representative (Trevor Jennings, Bloo Anderson).

## Abstract

**Background:** Stroke survivors rate longer-term (>2 years) psychological recovery as their top priority, but data on how frequently psychological consequences occur is lacking. Prevalence of cognitive impairment, depression/anxiety, fatigue, apathy and related psychological outcomes, and whether rates are stable in long-term stroke, is unknown.

**Methods:** *N* = 105 long-term stroke survivors (*M* [*SD*] age = 72.92 [13.01]; *M* [*SD*] acute NIH Stroke Severity Score = 7.39 [6.25]; 59.0% Male; *M* [*SD*] years post-stroke = 4.57 [2.12]) were recruited (potential *N* = 208). Participants completed 3 remote assessments, including a comprehensive neuropsychological battery, and questionnaires on emotional distress, fatigue, apathy and other psychological outcomes. Ninety participants were re-assessed one year later. Stability of outcomes was assessed by Cohen’s *d* effect size estimates and percent Minimal Clinically Important Difference changes between time points.

**Results:** On the Montreal Cognitive Assessment 65.3% scored <26. On the Oxford Cognitive Screen 45.9% had at least one cognitive impairment. Attention (27.1%) and executive function (40%) were most frequently impaired. 23.5% and 22.5% had elevated depression/anxiety respectively. Fatigue (51.4%) and apathy (40.5%) rates were high. Attention (*d* = −0.12; 85.8% stable) and depression (*d* = 0.09, 77.1% stable) were the most stable outcomes. Following alpha-adjustments, only perceptuomotor abilities (*d* = 0.69; 40.4% decline) and fatigue (*d* = −0.33; 37.2% decline) worsened over one year. Cognitive impairment, depression/anxiety, fatigue and apathy all correlated with worse quality of life.

**Conclusion:** Nearly half of participants >2 years post-event exhibited psychological difficulties, which impact long-term quality of life. Stroke is a chronic condition requiring long-term psychological support.

## Introduction

Globally, stroke is the second highest cause of mortality and is known to increase risk of chronic disability, sustained physical and cognitive impairment, and poorer quality of life that affects both the stroke survivor and carers^1–3^. The typical profile of long-term psychological outcomes post-stroke is not well-characterized, despite the increased likelihood of long-term impairments due to higher survival rates^4, 5^. In a systematic review of unmet care needs of stroke survivors^6^ managing psychological outcomes was the most frequently reported unmet need. Psychological information needs were shown to increase from 6 months (22.4%) to 2 years post-stroke (81.4%)^6^ due to requiring further information when initial recovery is made or receiving irrelevant information in early stroke.

Psychological outcomes of stroke can include poor attention, memory, executive function, perceptuomotor and language abilities^7^, mental health difficulties such as depression and anxiety^8, 9^, and extended outcomes such as fatigue and apathy^10, 11^. Difficulties in any of these in early stroke are known to contribute to poorer quality of life, and may reduce daily activity participation^12, 13^ and increase need for carer support^4^. However, despite the recognized long-term importance of psychological outcomes^14–16^, research has been mainly limited to the first year post-stroke^13, 17, 18^.

Longitudinal assessment of cognitive function after stroke has been predominantly completed with brief global screening tools such as the Mini-Mental State Examination (MMSE)^19, 20^. Though choice of tool will depend on the desired cognitive information, post-stroke cognitive impairment often includes deficits in a variety of domains^21^, and an in-depth neuropsychological assessment is more feasible outside of the acute window. Mild cognitive impairments are thought to be common in chronic stroke^22^ and sensitive neuropsychological assessment is therefore warranted in long-term follow-up to detect these often more subtle domain-specific impairments that can still impact quality of life^23^.

Emotional distress after stroke is also common, with post-stroke depression and anxiety estimated to affect 31.0% and 24.2% of stroke survivors respectively^8, 9^. These are frequently accompanied by additional outcomes that can have psychological causes, including fatigue^10^, apathy^11^ and poor sleep quality^24, 25^. Though evidence suggests these extended outcomes are more common in acute stroke, prevalence rates remain stable at 1-year^26, 27^. Cognitive impairment has been shown to double the risk for emotional distress and extended outcomes in the first-year post-stroke^28–30^ and strongly relates to long-term participation^31^. This emphasizes the need to understand the psychological consequences in a wide variety of areas and considered holistically with regards to their collective impact on post-stroke quality of life.

The temporal nature of these various psychological outcomes in chronic stroke is not well-described. Variability in emotional distress^32^ and cognitive functioning (heterogeneous patterns of improvement and decline^33^) has been examinded in early stroke. However, research in long-term stroke has focused mainly on cognitive decline and dementia diagnoses^17^. A more complete and improved understanding of the prevalence and nature of various long-term psychological outcomes is essential to tailoring community stroke services to the needs of stroke survivors.

## Study Aims and Objectives

The OX-CHRONIC study aimed to characterize the psychological profiles of long-term stroke (at least 2 years post-stroke). The primary objective was to identify the long-term prevalence of clinical impairment in six specific cognitive domains (language abilities, number processing, apraxia, memory, spatial attention, and executive function), and extended psychological consequences including depression, anxiety, fatigue, and apathy. Stability of psychological outcomes within a year’s time, and the impact of these psychological consequences on quality of life, was also examined.

## Methods

All participants provided informed consent to take part. The study was approved by NRES ethics committee (REC Reference: 19/SC/0520).

### Participants

Participants were recruited from the Oxford Cognitive Screening programme, a stroke cohort that had been consecutively recruited from the acute stroke ward within the John Radcliffe Hospital, UK between 2012 and 2020 (see protocol; Demeyere et al^34^). Participants who consented to future studies with the research team following a 6-month post-stroke assessment and who were at least 2 years post-stroke (*N* = 208) were contacted for participation in OX-CHRONIC. Participants consenting to OX-CHRONIC completed a battery of self-report and neuropsychological measures across two time points one year apart (termed Wave 1 and Wave 2), and optionally wore an activity monitor for one week following assessment^1^. With stroke participant consent, their carers were approached about participation, and carers consenting to participation completed self-report questionnaires. Due to the COVID-19 pandemic, all OX-CHRONIC assessments took place remotely either over the telephone or via videoconferencing in up to 3 separate sessions per time point. A detailed description of the full study protocol is reported elsewhere^34^.

### Study Measures

Neuropsychological assessments selected were based on their wide-range use in stroke settings and covered a wide range of possible cognitive domain impairments. This included domain-general cognition (MoCA^35^), stroke-specific cognition (Oxford Cognitive Screen [OCS]^21^) language (Cookie Theft Task^36^; Boston Naming Test^37^; Letter and Category Fluency^38^), executive function (Trail Making Test A & B^38^; Hayling Sentence Completion Test^39^; OCS-Plus Mixed Trails^40^), memory (Digit Span Forwards & Backwards^41^; Logical Memory Test^41^; Picture Memory Test^40^), attention (Star Cancellation Test^42^), and perceptuomotor abilities (OCS-Plus Figure Copy Test^40^; Rey-Osterrieth Complex Figure Copy Test^43^).

Self-report questionnaires were similarly selected across a range of psychological outcomes (e.g., subjective cognition, emotional distress) and functional information (e.g., activities of daily living). This included measures of cognitive abilities (Cognitive Failures Questionnaire^44^; Cognitive Reserve Index^45^), daily function (Telephone Modified Rankin Scale^46^; Nottingham Extended Activities of Daily Living Scale^47^; 3-item Barthel Index^48^), emotional distress (Hospital Anxiety and Depression Scale [HADS]^49^; Geriatric Depression Scale [GDS]^50^), extended outcomes such as fatigue (Fatigue Severity Scale [FSS]^51^), apathy (Apathy Evaluation Scale [AES]^52^) and sleep quality (Sleep Condition Indicator-8 [SCI-8]^53^), and quality of life measures (EQ-5D-5L^54^; Stroke Impact Scale-Short Form [SF-SIS]^55^; World Health Organization Quality of Life Scale^56^; ICEpop Capability Measure for Adults^57^). Carer measures included the Caregiver Strain Index^58^, the Informant-GDS^59^, and the Informant Questionnaire for Cognitive Decline in the Elderly (IQ-CODE)^60^. An overview of study measures is in *Supplementary Table 1* and in the study protocol^34^.

### Statistical Analyses

Analyses were performed using R version 4.2.1^61^. The datasets analysed and code for the current study are available at osf.io/y2mev

Descriptive statistics of Wave 1 and Wave 2 study variables were calculated. Where available per measure, validated cut scores (binarized as yes/no) were used to determine percentage of participants with cognitive impairment (for neuropsychological assessments) and scores that indicate elevated symptoms/functional difficulties warranting clinical attention (collectively termed “clinically significant” within the manuscript; for self-report questionnaires only). For study measures, cut scores were developed based on comparison to normative data in healthy adults or based on sensitivity/specificity analyses. Cut scores used in the present study are shown in *Supplementary Table 1.* 95% confidence intervals for percent estimates were calculated using the below formula:

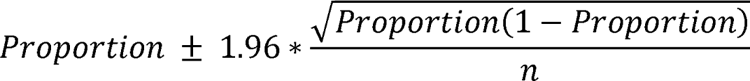

To account for potential risk or increased rates of impairment across the large number of more sensitive in-depth neuropsychological measures, chi-square tests with false discovery rate corrections were used to determine whether the proportion of those impaired versus not impaired at each time point differed (see *Supplementary Materials*). Additionally, we do not present data on the proportion of participants with any impairment on the in-depth neuropsychological assessments to further reduce this risk.

To determine stability of psychological outcomes, paired t-tests (for parametric data) and Wilcoxon rank-sum tests (for non-parametric data) were used to determine whether a statistically significant change occurred on study measures (instead of proportion of those meeting cut score criteria above) between Wave 1 and Wave 2. Family-wise alpha corrections across neuropsychological assessments and self-report measures were alpha-adjusted using false discovery rate (FDR) corrections to balance between risk of Type I and Type II errors. Cohen’s *d* was additionally estimated to measure effect size differences. As a comparator, distribution-based Minimal Clinically Important Difference (MCID) estimates (i.e., 0.5 standard deviation change) were used to determine the percentage of participants whose scores were of clinical relevance from Wave 1 to Wave 2. This approach was taken given that some OX-CHRONIC measures do not have published MCIDs in stroke (e.g., Hayling Sentence Completion Test). Where available in the literature per measure, anchor-based MCIDs were additionally used to determine clinically relevant change.

To examine whether potential differences existed at Wave 1 from those retained versus those lost to attrition at Wave 2, independent t-tests were conducted comparing demographics (age, sex, handedness, years of education, stroke type, stroke severity, years post-stroke), cognitive impairment (OCS language, memory, attention, number processing, and executive function impairments), and stroke related quality of life scores (SF-SIS stroke recovery score, hand function, arm function, mobility, activities of daily living, emotions, communication, memory and participation). Results are in the *Supplementary Materials*.

To explore the impact of psychological outcomes on quality of life, Spearman rank correlations were conducted between cognition (MoCA), depression/anxiety (HADS), fatigue (FSS) and apathy (AES) to EQ-5D-5L health rating scores and SF-SIS scores at Wave 1.

## Results

A total of 105 stroke participants completed OX-CHRONIC Wave 1, with 90 completing re-assessment at Wave 2 one year later. Seventy-four carers participated in Wave 1, and 66 in Wave 2. A recruitment flow chart is shown in Figure 1 (see study protocol for further details on study sample eligibility^34^)

**Figure 1.**
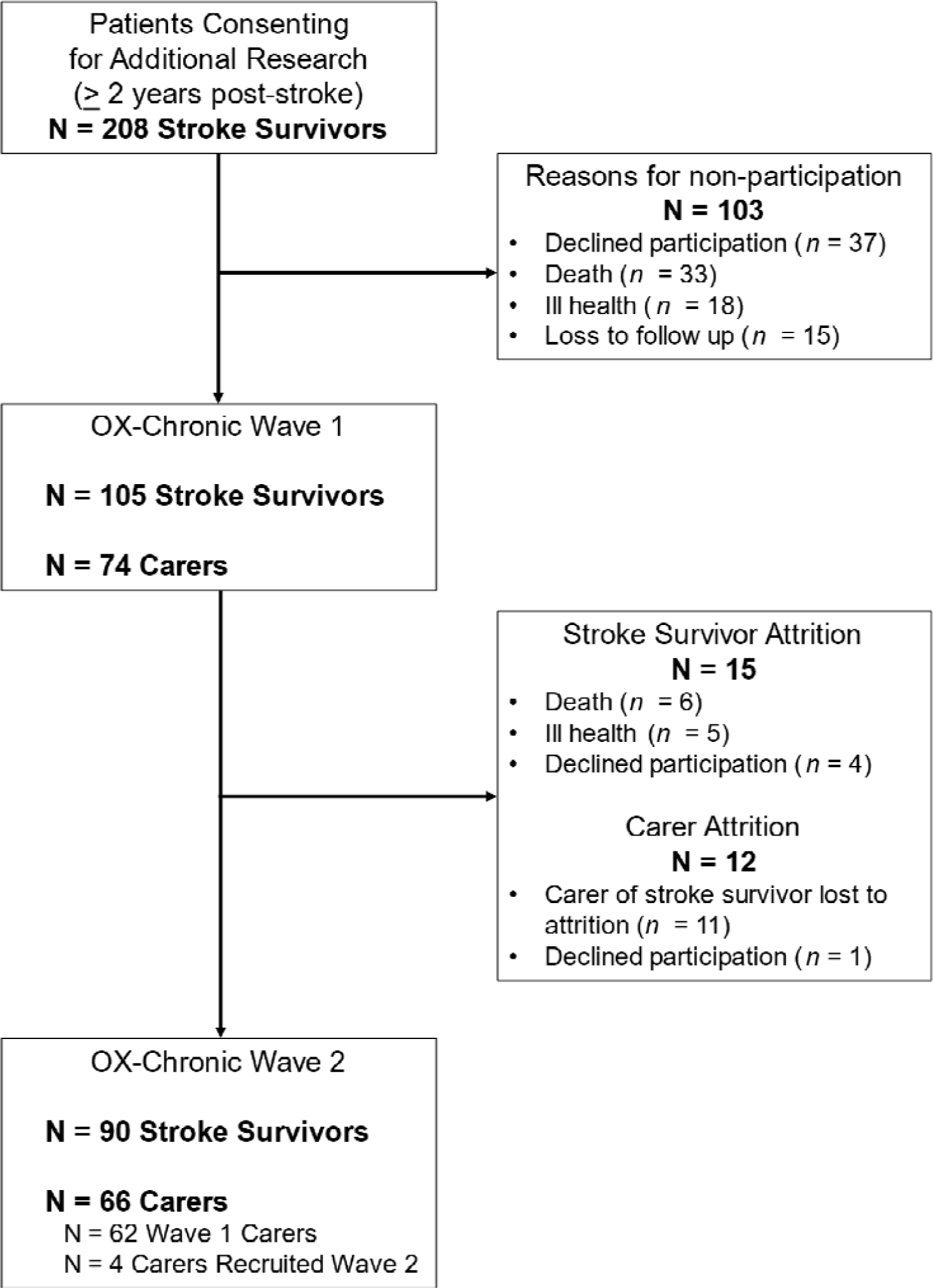
Participant recruitment flowchart to OX-CHRONIC at Wave 1 and Wave 2.

Participant demographics are presented in Table 1. Our cohort included a high proportion of individuals with left hemisphere stroke (40.00%) and moderate stroke severity scores (median NIHSS = 5).

**Table 1.** Participant Characteristics. NIHSS = National Institute of Health Stroke Severity

| **Participants (N = 105)** |  | Min - Max |
| --- | --- | --- |
| **Sex – n (%)** |  |  |
| Male | 62 (59.05%) |  |
| Female | 43 (40.95%) |  |
| **Age – Mean (SD)** | 72.92 (13.01) | 21 – 96 |
| **Years Post-Stroke – Mean (SD)** | 4.57 (2.12) | 2 – 9.38 |
| **Years of Education – Mean (SD)** | 13.94 (3.67) | 9 – 23 |
| **Stroke Type – n (%)** |  |  |
| Ischaemic | 88 (83.80%) |  |
| Haemorrhagic | 17 (16.20%) |  |
| **Lesion Hemisphere – n (%)** |  |  |
| Left | 42 (40.00%) |  |
| Right | 41 (39.05%) |  |
| Bilateral | 8 (7.60%) |  |
| Undetermined from scan | 14 (13.35%) |  |
| **First or Recurrent Stroke – n (%)** |  |  |
| First | 70 (66.67%) |  |
| Recurrent | 35 (33.33%) |  |
| **Acute NIHSS Score – Mean (SD)** | 7.39 (6.25) | 0 – 27 |
| **Carers (N = 74)** |  |  |
| **Sex – n (%)** |  |  |
| Male | 27 (36.50%) |  |
| Female | 47 (63.50%) |  |
| **Relationship to Participant – n (%)** |  |  |
| Wife | 35 (47.30%) |  |
| Husband | 24 (32.40%) |  |
| Daughter/Son | 7 (9.40%) |  |
| Parent | 5 (6.80%) |  |
| Other | 3 (4.10%) |  |

### Participant Attrition and Study Outcomes

Differences in demographics and study measures between those retained (*N* = 90) and those lost to attrition at Wave 2 (*N* = 15) are reported in *Supplementary Table 2.* Overall, there were no significant differences in demographics or cognition. However, participants lost to attrition self-reported worse overall SF-SIS functioning, lower levels of activities of daily living (ADLs), and worse emotional distress at Wave 1. When comparing those lost to attrition not due to death (*N* = 9) and those retained, there were no statistically significant difference in any variables examined.

### Chronic Cognitive Impairment

Full details of impairment frequency per neuropsychological measure, per domain, is shown in Table 2. Detailed descriptive statistics (i.e., minimum and maximum scores, task times) are in *Supplementary Tables 3* and *4*.

**Table 2.** Descriptive statistics and percent prevalence of impairment status on the stroke-specific Oxford Cognitive Screen subtasks (italicized) and in-depth neuropsychological assessments per domain at Wave 1 (*N* = 98) and Wave 2 (*N* = 85) with 95% confidence intervals. Impairment scores are determined based on comparison to normative data in healthy adults. OCS: Oxford Cognitive Screen; MoCA: Montreal Cognitive Assessment; WMS: Wechsler Memory Scale; BIT: Behavioural Inattention Test; ROCF: Rey-Osterrieth Complex Figure

| DOMAIN | WAVE 1 (N = 98) |  | WAVE 2 (N = 85) |  |
| --- | --- | --- | --- | --- |
|  | Mean (SD) | Impairment Status<br>N (% [95% CI]) | Mean (SD) | Impairment Status<br>N (% [95% CI]) |
| **Domain-General Cognition** |  |  |  |  |
| *OCS Tasks Impaired* | 0.90 (1.40) | 45 (45.92 [36.1 – 55.8]) | 0.99 (1.33) | 40 (47.06 [36.5 – 57.7]) |
| MoCA (<26) | 23.56 (4.16) | 64 (65.31 [55.9 – 74.7]) | 22.98 (4.51) | 57 (67.06 [57.1 – 77.1]) |
| MoCA (<22) | -- | 30 (30.61 [21.5 – 39.7]) | -- | 29 (34.12 [24.0 – 44.2]) |
| **Language** |  |  |  |  |
| *OCS Picture Naming* | 3.65 (0.59) | 4 (4.1 [0.2 – 7.9]) | 3.71 (0.53) | 3 (3.5 [-0.4 – 7.5]) |
| *OCS Semantics* | 3 (0) | 0 (0.0 [0 – 0]) | 2.99 (0.11) | 1 (1.2 [-1.1 – 3.5]) |
| *OCS Sentence Reading* | 14.62 (1.56) | 7 (7.1 [2.0 – 12.2]) | 14.59 (1.19) | 9 (10.6 [4.0 – 17.1]) |
| Cookie Theft Complexity | 0.81 (0.23) | 1 (1.0 [-0.9 – 3.1]) | 1.22 (0.24) | 1 (1.2 [-1.1 – 3.5]) |
| Boston Naming Test | 13.89 (1.79) | 4 (4.1 [0.2 – 7.9]) | -- | -- |
| Letter Fluency Total | 32.53 (14.94) | 14 (14.3 [7.4 – 21.2]) | 32.69 (14.88) | 9 (10.7 [4.1 – 17.3]) |
| Category Fluency Total | 31.64 (10.00) | 8 (8.2 [2.7 – 13.6]) | 32.51 (11.09) | 7 (8.3 [2.5 – 14.2]) |
| **Executive Function** |  |  |  |  |
| *OCS Mixed Trails* | 10.78 (3.64) | 14 (14.3 [7.4 – 21.2]) | 11.61 (2.17) | 4 (4.7 [0.2 – 9.2]) |
| Trails A Accuracy | 23.81 (0.63) | 11 (11.3 [5.1 – 17.6]) | 23.48 (1.89) | 8 (9.4 [3.2 – 15.6]) |
| Trails B Accuracy | 18.54 (6.00) | 28 (28.9 [19.9 – 37.8]) | 18.04 (5.55) | 34 (40.0 [29.6 – 50.4]) |
| Hayling Test Total | 12.03 (3.65) | 30 (30.6 [21.5 – 39.7]) | 13.60 (3.93) | 20 (23.5 [14.5 – 32.6]) |
| OCS-Plus Mixed Trails | 10.55 (4.16) | 15 (15.5 [8.3 – 22.6]) | 10.61 (3.76) | 23 (27.1 [17.6 – 36.5]) |
| **Memory** |  |  |  |  |
| *Orientation* | 3.93 (0.39) | 4 (4.1 [0.2 – 7.9]) | 3.91 (0.40) | 5 (5.9 [0.9 – 10.9]) |
| *Recall* | 2.57 (1.32) | -- | 2.74 (1.14) | -- |
| *Recall + Recognition* | 3.71 (0.70) | 5 (5.1 [0.8 – 9.5]) | 3.76 (0.55) | 3 (3.5 [-0.4 – 7.5]) |
| *Episodic Recognition* | 3.84 (0.40) | 1 (1.0 [-0.9 – 3.0]) | 3.79 (0.41) | 0 (0.0 [0 – 0]) |
| Digit Span Forwards | 7.49 (2.33) | 9 (9.3 [3.5 – 15.0]) | 7.25 (2.57) | 7 (8.2 [2.4 – 14.1]) |
| Digit Span Backwards | 5.90 (2.14) | 4 (4.1 [0.2 – 7.9]) | 5.89 (2.36) | 5 (5.8 [0.8 – 10.9]) |
| Logical Memory I | 12.37 (4.46) | 3 (3.1 [-0.3 – 6.5]) | 12.24 (4.50) | 2 (2.4 [-0.8 – 5.6]) |
| Logical Memory II | 10.67 (5.19) | 9 (9.3 [3.5 – 15.0]) | 10.73 (4.96) | 6 (7.1 [1.6 – 12.5]) |
| Picture Memory Test | 9.93 (2.73) | 26 (26.5 [17.8 – 35.3]) | -- | -- |
| **Visuospatial Attention** |  |  |  |  |
| *Broken Hearts Accuracy* | 43.53 (7.88) | 21 (21.4 [13.3 – 29.6]) | 43.76 (6.57) | 23 (27.1 [17.6 – 36.5]) |
| *Broken Hearts Time* | 126.73 (38.31) | -- | 130.33 (37.05) | -- |
| *Egocentric Neglect* | -- | 8 (8.2 [2.7 – 13.6]) | -- | 7 (8.2 [2.4 – 14.1]) |
| *Allocentric Neglect* | -- | 8 (8.2 [2.7 – 13.6]) | -- | 7 (8.2 [2.4 – 14.1]) |
| Star Cancellation Total | 52.19 (5.70) | 18 (18.6 [10.9 – 26.3]) | 52.76 (2.26) | 14 (16.5 [8.6 – 24.4]) |
| **Number Processing** |  |  |  |  |
| *OCS Number Writing* | 2.85 (0.41) | 12 (12.2 [5.8 – 18.7]) | 2.79 (0.56) | 14 (16.5 [8.6 – 24.4]) |
| *OCS Calculation* | 3.77 (0.49) | 3 (3.1 [-0.4 – 6.5]) | 3.69 (0.51) | 4 (4.7 [0.2 – 9.2]) |
| **Perceptuomotor Abilities** |  |  |  |  |
| OCS-Plus Figure Copy | 54.99 (5.85) | 6 (6.1 [1.4 – 10.9]) | 54.07 (5.19) | 5 (5.9 [0.9 – 10.9]) |
| OCS-Plus Figure Recall | 41.46 (10.57) | 6 (6.1 [1.4 – 10.9]) | 39.81 (9.65) | 9 (10.6 [4.1 – 17.1]) |
| ROCF Copy | 26.68 (6.38) | 10 (10.2 [4.2 – 16.2]) | 24.04 (6.35) | 17 (20.2 [11.7 – 28.8]) |
| ROCF Recall | 12.79 (7.35) | 8 (8.3 [2.9 – 13.8]) | 12.42 (6.61) | 4 (9.4 [3.2 – 15.6]) |

At Wave 1, the majority of participants (65.3%) were classified as having a domain-general cognitive impairment on the MoCA (score <26). When using a stroke-specific, multidomain cognitive impairment cutoff score of 22^62^, prevalence of impairment was one-third of the sample at both time points (30.6% Wave 1; 34.1% Wave 2).

At Wave 1, 45.9% (*N* = 45) of participants were impaired on at least one of the 10 OCS subtasks (i.e., scored below normative performance of healthy controls^21^). At Wave 2, 47.0% (*N* = 40) of participants were impaired in at least one subtask. There was no significant difference between Wave 1 and Wave 2 in presence of any OCS domain impairment (χ^2^ = 0.24, *p* = 0.88), and average number of OCS subtasks impaired (*F*_1,181_ = 0.20, *p* = 0.66).

Across assessment timepoints, attention impairments, particularly in selective visual attention rather than visuospatial neglect, were the most frequently observed using the OCS (Wave 1 21.4% [95% CI 13.3 – 29.6]). When using in-depth neuropsychological measures executive function impairments were most prevalent (Wave 1 30.6% [95% CI 21.5 – 39.7]). Participants were least likely to have expressive language deficits (as low as 1.0% on discourse language on the Cookie Theft task). Notably, participants performed well on verbal working memory and verbal episodic recall tasks (e.g., Digit Span Backward, Logical Memory Immediate Recall; impairment rates of 4.1% and 5.1% respectively), while a comparatively high proportion were impaired on the Picture Memory Test (26.5%). In investigating whether proportion of those impaired changed across time points, we find that proportion of impairment stayed stable (see *Supplementary Table 5*).

### Self-reported Emotional Distress, Extended Outcomes, and Quality of Life in Chronic Stroke

Descriptive statistics of questionnaire data are shown in Table 3. Detailed descriptive statistics (i.e., subscale scores, ranges) are shown in *Supplementary Table 6*.

**Table 3.** Descriptive statistics and proportion of sample with clinically elevated scores on self-reported questionnaires for participants and carer-reported measures with 95% confidence intervals. Cut scores used were taken from each scales’ published psychometric analysis were used to indicate percent of participants with elevated symptoms or scores warranting clinical attention (termed “clinically significant” within the table). SIS: Stroke Impact Scale; WHO-QoL-BREF: World Health Organization Quality of Life Scale – Abbreviated; ADL: Activities of Daily Living; ICECAP-A: ICEpop Capability Measure for Adults; EQ5D-5L: EuroQol-5 Dimensions-5 Levels; CFQ: Cognitive Failures Questionnaire; HADS: Hospital Anxiety and Depression Scale; GDS: Geriatric Depression Scale; CSI: Caregiver Strain Index; IQ-CODE: Informant Questionnaire for Cognitive Decline in the Elderly ^1^Data presented from Wave 1 ^2^Data presented from Wave 2

| Domain | N | Mean (SD) | Clinically Significant –<br>N (% [95% CI]) |
| --- | --- | --- | --- |
| **Post-Stroke Abilities** |  |  |  |
| Modified Rankin Scale Score<sup>1</sup> | 100 | 1.79 (1.27) | 30 (28.6 [19.7 – 37.4]) |
| Barthel-3 Item Short Form Total<sup>1</sup> | 99 | 6.93 (1.58) | -- |
| Nottingham Extended ADL<sup>1</sup> | 100 | 47.77 (16.26) | -- |
| **Quality of Life** |  |  |  |
| SIS-Short Form Scaled Total<sup>1</sup> | 102 | 72.66 (20.12) | -- |
| SIS-Long Form Stroke Recovery Score<sup>2</sup> | 88 | 71.91 (21.63) | -- |
| WHO-QoL-BREF Overall Score<sup>2</sup> | 83 | 7.22 (1.65) | -- |
| ICECAP-A Total<sup>1</sup> | 101 | 15.65 (2.90) | -- |
| EQ5D-5L Health Rating<sup>1</sup> | 99 | 68.61 (18.83) | -- |
| **Cognitive Ability** |  |  |  |
| CFQ Total<sup>2</sup> | 88 | 33.60 (17.93) | 29 (32.2 [22.5 – 41.9]) |
| Cognitive Reserve Index<sup>2</sup> | 84 | 128.42 (19.68) | -- |
| **Emotional Distress** |  |  |  |
| HADS-Depression Total<sup>1</sup> | 98 | 4.97 (3.98) | 23 (23.5 [15.1 – 31.9]) |
| HADS-Anxiety Total<sup>1</sup> | 98 | 5.23 (4.02) | 22 (22.5 [14.2 – 30.7]) |
| GDS Total<sup>1</sup> | 101 | 4.22 (4.10) | 34 (33.7 [24.4 – 42.9]) |
| **Extended Outcomes** |  |  |  |
| Apathy Evaluation Scale<sup>1</sup> | 101 | 32.36 (10.21) | 41 (40.6 [31.0 – 50.2]) |
| Fatigue Severity Scale<sup>1</sup> | 101 | 35.56 (15.34) | 52 (51.5 [41.7 – 61.2]) |
| Sleep Condition Indicator<sup>1</sup> | 100 | 23.29 (8.04) | 21 (21.0 [13.0 – 28.9]) |
| **Carer Measures** |  |  |  |
| CSI Total Score<sup>1</sup> | 68 | 2.76 (3.12) | 9 (13.2 [5.2 – 21.2]) |
| Informant-GDS Total<sup>1</sup> | 70 | 5.18 (4.30) | 35 (50.0 [38.2 – 61.7]) |
| IQ-CODE<sup>1</sup> | 74 | 3.23 (0.63) | 29 (39.2 [28.1 – 50.3]) |

Though 30.0% reported at least a moderate disability on the Modified Rankin Scale (score >3), 55% had a slight disability that affected performance on daily activities (score >2). Prevalence of self-reported cognitive difficulties were overall lower than that observed using objective neuropsychological measures, with 32.2% reporting clinically significant levels of cognitive failures in everyday life. However, 40% of carers rated their stroke survivor relative at risk of cognitive decline. Prevalence rates for emotional distress varied by measure – 23.5% and 22.5% of stroke survivors reported mild depression and mild anxiety respectively on the HADS, lower than GDS rates (33.4%). Further, 50% of carers rated their relative as having at least mild depression on the informant GDS^38^. Extended outcomes were more frequently endorsed, with clinically significant rates of fatigue observed in 51.5% of participants, and high rates of apathy (40.6%). Significant sleep difficulties were the least frequently reported outcome by stroke survivors (21.0%).

Despite moderate levels of emotional distress and extended outcomes, EQ-5D-5L quality of life scores were comparable to healthy population norms in a similar age bracket^63^, and stroke-related quality of life was moderate. Significant carer strain was also low (13.2%).

### Stability of Psychological Outcomes between Wave 1 and Wave 2

An overview of whether change in outcomes was statistically and/or clinically meaningful between time points is in Table 4. Detailed information of stability (e.g., comparison to anchor-based MCIDs; test statistics) is shown in *Supplementary Table 7*.

**Table 4.** Stability results of neuropsychological assessment, self-report, and carer measures per domain between Wave 1 and Wave 2 including Cohen’s *d* effect size estimates. Statistical tests were alpha-adjusted using family-wise false discovery rate (FDR) corrections. Distribution-based MCIDs were estimated by calculating percentage of individuals whose difference in scores per measure between Wave 1 and Wave 2 were 0.5 standard deviations (SDs) above or below the mean of each measure at Wave 1. MCID: Minimal Clinically Important Difference; OCS: Oxford Cognitive Screen; ROCF: Rey-Osterrieth Complex Figure Copy; SIS: Stroke Impact Scale; EQ5D-5L: EuroQol-5 Dimensions-5 Levels; HADS: Hospital Anxiety and Depression Scale; GDS: Geriatric Depression Scale; IQ-CODE: Informant Questionnaire for Cognitive Decline in the Elderly * *p* < 0.05, ** *p* < 0.01, *** *p* < 0.001 following family-wise FDR corrections.

| Domain | Cohen's $d$ | MCID<br>(0.5 SD) | Distribution MCID | | |
| --- | --- | --- | --- | --- | --- |
|  |  |  | Improve | Decline | No Change |
| **Domain-General Cognition** |  |  |  |  |  |
| OCS Tasks Impaired | -0.10 | 0.69 | 21.2% | 24.7% | 54.1% |
| Montreal Cognitive Assessment | 0.28 | 2.07 | 11.7% | 28.2% | 60.1% |
| **Language** |  |  |  |  |  |
| Cookie Theft Complexity | **-1.33\*\*\*** | 0.12 | **83.1%** | 3.6% | 13.3% |
| Letter Fluency Total | 0.17 | 7.46 | 8.2% | 17.6% | 74.2% |
| Category Fluency Total | -0.04 | 5.00 | 16.4% | 15.3% | 68.3% |
| **Executive Function** |  |  |  |  |  |
| Trail Making Test A Accuracy | 0.17 | 0.32 | 7.1% | 15.4% | 77.5% |
| Trail Making Test B Accuracy | 0.14 | 2.99 | 20.2% | 27.3% | 52.5% |
| Hayling A Response Time | **0.46\*\*\*** | 14.52 | **32.1%** | 4.7% | 63.2% |
| Hayling B Response Time | 0.28 | 23.98 | 36.9% | 8.3% | 54.8% |
| Hayling B Errors | -0.04 | 5.67 | 24.1% | 28.9% | 47.0% |
| Hayling Test Total | **-0.49\*\*\*** | 0.92 | **60.2%** | 25.3% | 14.5% |
| OCS-Plus Mixed Accuracy | 0.01 | 2.08 | 17.8% | 21.4% | 60.8% |
| **Memory** |  |  |  |  |  |
| Digit Span Forwards | 0.12 | 1.16 | 3.5% | 49.4% | 47.1% |
| Digit Span Backwards | 0.04 | 1.07 | 18.8% | 20.0% | 61.2% |
| Logical Memory Immediate | 0.09 | 2.23 | 23.5% | 30.9% | 45.6% |
| Logical Memory Recall | 0.03 | 2.59 | 23.8% | 29.7% | 46.5% |
| **Visuospatial Attention** |  |  |  |  |  |
| Star Cancellation Task Total | -0.12 | 2.85 | 9.5% | 4.7% | 85.8% |
| **Perceptuomotor Abilities** |  |  |  |  |  |
| OCS-Plus Figure Copy | 0.17 | 2.92 | 12.9% | 31.7% | 55.4% |
| OCS-Plus Figure Recall | 0.22 | 5.28 | 17.6% | 31.7% | 50.7% |
| ROCF – Copy | **0.69\*\*\*** | 3.19 | 3.5% | **40.4%** | 56.1% |
| ROCF – Recall | 0.17 | 3.67 | 19.2% | 22.8% | 58.0% |
| **Post-Stroke Abilities** |  |  |  |  |  |
| Modified Rankin Scale Score | -0.23 | 0.63 | 16.4% | 29.4% | 54.2% |
| Barthel-3 item Total | -0.12 | 0.79 | 24.7% | 14.1% | 61.2% |
| **Quality of Life** |  |  |  |  |  |
| SIS-Short Form Scaled Total | -0.05 | 10.05 | 1.1% | 1.1% | 97.8% |
| EQ5D-5L Health Rating | **-0.29\*** | 9.41 | **37.2%** | 16.2% | 46.6% |
| **Emotional Distress** |  |  |  |  |  |
| HADS-Depression Total | -0.08 | 1.99 | 24.7% | 25.8% | 49.5% |
| HADS-Anxiety Total | -0.13 | 2.01 | 12.9% | 21.1% | 66.0% |
| GDS Total | 0.09 | 2.04 | 16.1% | 6.9% | 77.1% |
| **Extended Outcomes** |  |  |  |  |  |
| Apathy Evaluation Scale | 0.06 | 5.10 | 12.7% | 14.9% | 72.4% |
| Fatigue Severity Scale | **-0.33\*** | 7.66 | 15.1% | **37.2%** | 47.7% |
| Sleep Condition Indicator | 0.07 | 4.02 | 11.6% | 12.7% | 75.7% |
| **Carer Measures** |  |  |  |  |  |
| Informant-GDS Total | -0.10 | 2.15 | 9.1% | 9.1% | 81.8% |
| IQ-CODE | 0.34 | 0.31 | 15.5% | 5.1% | 79.4% |

From a statistical perspective, domain-general cognition remained stable between years as measured by number of OCS subtasks impaired (Wilcoxon’s *V* = 403.50, *p* = 0.39, Cohen’s *d* = −0.10) and MoCA scores (*t* = 2.57, *p* = 0.053, *d* =0.28). However, when considering anchor-based MCID change, 42.3% of stroke survivors showed decline on the MoCA (vs. 28.2% distribution-based). In an exploratory analysis, visuospatial scores were the only MoCA subtests to decline (*t* = 2.52, *p* = 0.01).

Memory (*d*s = 0.03 – 0.12) and visuospatial attention tasks (*d* = −0.12) had negligible effect size differences between Wave 1 and Wave 2, though we note 49.4% of participants showed MCID decline on verbal memory on the Digit Span forwards. Discourse language (*d = -*1.33; 83.1% MCID improvement) and executive function tasks (*d = -*0.49, 60.2% MCID improvement) demonstrated moderate to large improvements between years, though complex figure copy abilities showed moderate decline (*d =* 0.69; 40.4% MCID decline).

Regarding self-report and carer questionnaires, there were negligible effect size differences across all domains; with emotional distress measures remaining the most stable (*d*s = −0.10 – 0.09; MCID no change = 49.5% – 81.8%). However overall perceptions of health (EQ-5D-5L Health Ratings) improved between years (*d* = −0.29, MCID improvement = 37.2%), while fatigue worsened over time (*d* = −0.33, MCID decline = 37.2%).

Notably, even in this long-term stroke cohort, some measures showed MCID improvement between years – for example, 36.9% had improved executive function abilities and 24.7% had improved depression.

### Impact of Psychological Outcomes on Quality of Life

Median participant scores were moderate across all SIS domains, though considerable variation was present (see *Supplementary Figure 1*). Scaled scores were highest in communication (median = 89.29) and lowest in emotions (median = 72.22).

Scatter plots of cognition, depression, anxiety, fatigue and apathy to long-term OX-CHRONIC quality of life measures is shown in Figure 2. Domain-general cognition as measured by the OCS and MoCA did not seem to impact on overall quality of life as measured by the EQ-5D-5L (*rho*s = −0.05 and 0.14 respectively); however worse cognitive outcomes correlated with worse stroke-specific quality of life (*rho*s = −0.23 and 0.34). In contrast, greater fatigue, depression, anxiety, and apathy all significantly correlated with worse overall quality of life on the EQ-5D-5L and the stroke-specific SF-SIS.

**Figure 2.**
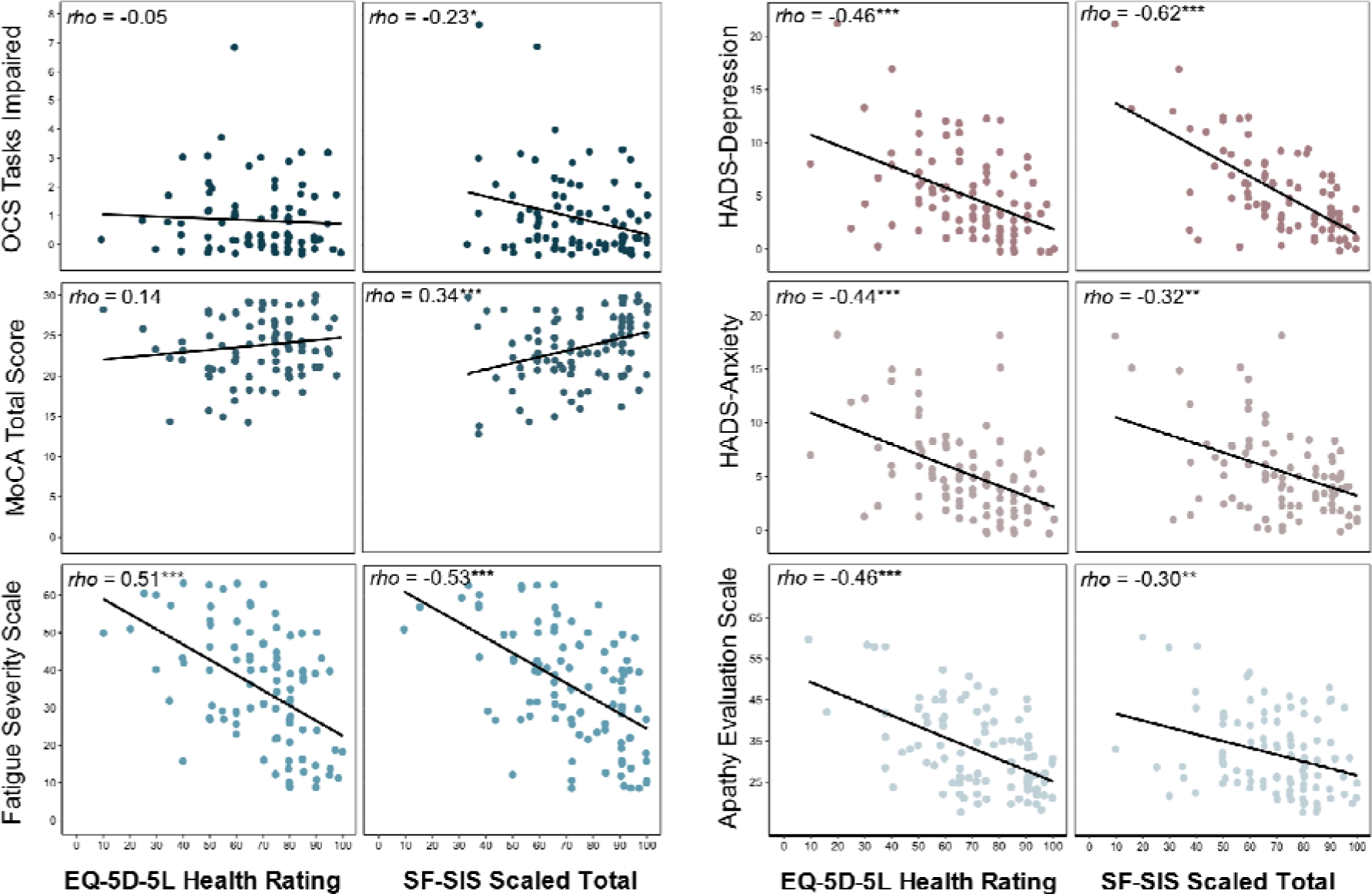
Scatter plots of measures of cognition (OCS, MoCA), depression (HADS-Depression), anxiety (HADS-Anxiety), fatigue (Fatigue Severity Scale) and apathy (Apathy Evaluation Scale) to overall quality of life and stroke-related quality of life using Wave 1 data. EQ-5D-5L: EuroQoL 5D-5L; SF-SIS: Short Form Stroke Impact Scale; OCS: Oxford Cognitive Screen; MoCA: Montreal Cognitive Assessment; HADS: Hospital Anxiety and Depression Scale * *p* < 0.05, ** *p* < 0.01, *** *p* < 0.001

## Discussion

This study is one of the first in-depth examinations of psychological outcomes in chronic stroke, including addressing long-term cognition, emotional distress, fatigue, and apathy. At an average of 4.5 years post-event, cognitive impairments were present in nearly half of all chronic stroke survivors. Mild to severe levels of depression and anxiety were present in 20% - 50% of stroke survivors. Of all outcomes, clinically significant fatigue was the most prevalent, occurring in just over half of participants. Over a one-year period, only perceptuomotor abilities and fatigue statistically worsened in this chronic sample, while all outcomes showed some clinically meaningful improvement. Lastly, improved psychological outcomes significantly correlated with better perceived quality of life.

### Prevalence of Domain-General Cognitive Impairment

Domain-general impairments, as measured by two brief screening tools, ranged from 30% (MoCA <22^62^) to 45% (OCS) to 65% (MoCA <26). Previous research has similarly highlighted wide-ranging estimates of domain-general cognitive impairment. In a London registry study 22% were estimated to have mild cognitive impairment at 5-years post-stroke on the MMSE^71^, whilst other studies report 84% to have mild cognitive impairment at 4 years post-stroke^72^ (MoCA <26). Other MoCA prevalence estimates have ranged from ∼79% at 3 years post-stroke^73^, ∼46% at 5 years post-stroke^74^, to ∼61% at 10 years post-stroke^75^. Over a one-year period in the present chronic stroke sample, prevalence rates of domain-general impairments were found to be fairly stable on the OCS (47%) and MoCA (<26; 67%). In meta-analyses of chronic post-stroke cognitive impairment, it would be valuable to assess whether the differences in reported prevalence rates is due to measurement error, stroke-specific vs generic screens, or demographic and clinical factors in the sample. Notably, self-report and carer measures estimated differing rates of cognitive impairment (32% and 39% respectively), demonstrating discrepancies between observed and perceived, subjective cognitive impairments. These are also valuable to consider in prevalence rates of domain-general cognition.

### Prevalence of Domain-Specific Cognitive Impairment

Domain-specific impairment rates in this cohort are similar to previous cohorts^76–78^. Estimates of domain-specific cognitive impairments varied between OCS brief screening tasks and in-depth neuropsychological assessments. Executive function impairments were the most prevalent using in-depth, sensitive neuropsychological assessments, whereas visuospatial attention impairments were the most prevalent on the OCS (21%), though not notably higher than the in-depth visuospatial assessment (19%). Verbal memory impairments were comparable across brief and in-depth assessments (range 4% - 9%). Visual memory impairments were observed in 27% of participants (higher than previous estimates of ∼5% at 2 years post-stroke^78^ and ∼10% at 7 months post-stroke^79^). Low rates of language impairment were observed in both brief and in-depth assessments (0% - 7%). Language fluency tests had higher rates of impairment (8% – 14%), possibly reflecting the additional executive demands needed for fluency tests. This difference between brief tests and in-depth assessments confirms that unless more sensitive neuropsychological tests are used, these more subtle impairments are likely to be missed in typical post-stroke care. Collectively, findings show executive function abilities, visual memory, and visuospatial attention may be particularly important to monitor in long-term stroke.

### Prevalence of Emotional Distress and Extended Outcomes

Depression and anxiety rates in this cohort (∼25%) are similar to reported estimates in early stroke of up to 12 months (22% anxiety^8^; 31% depression^9^), and in other chronic samples estimating depression at 15 years post-stroke (31%^80^). Notably, depression prevalence was higher when rated by carers (50%), replicating previous research highlighting discrepancies in early stroke survivor-proxy reports.^81^ Individuals may feel stigmatized about endorsing depression and minimize emotional impact of the stroke itself, thus carer responses may be more representative. However, carer ratings of participant depression may indicate concern for the stroke survivor, or reflect carer mood, thus stroke survivor reports may be more accurate.

Clinically significant fatigue was reported by 51% of participants, consistent with community-based stroke survivors estimates (range 38% - 68%^82^), meta-analyses (50%^10^), and early stroke fatigue rates (50%^83^). Our cohort had higher levels of apathy (41%) compared to a systematic review^84^ pooled prevalence estimate (35%) and milder stroke cohorts (∼36%^85^). Long-term stroke survivors may require improved intervention and support in these areas; however, fatigue and apathy may be more resistant to change relative to depression and anxiety^77^. Sleep difficulties in this cohort (21%) were less prevalent than meta-analytic estimates (38%^86^). Increases in daytime sleepiness are associated with greater time post-stroke, rather than difficulties falling or staying sleep^86^ and thus exploring how different sleep difficulties categorizations relate to function would be valuable.

Despite the high frequency of depression, anxiety, apathy, fatigue and sleep disturbances, significant carer strain was relatively low in this cohort (13.2%). Previous work has reported approximately 30% of carers experience significant strain at 6 months post-stroke^87^ and 42% at 12 months^88^. Beyond 12 months, carers may become more adept at coping with care responsibilities, or perhaps stroke survivors continue to restore capabilities and require less care. Further research could explore how carer strain changes in relation to care competency and functional capability of the stroke survivor beyond 12 months. Irrespective of carer strain, a systematic review of long-term unmet needs of carers (up to 4 years post-stroke) showed the need for continued psychological information and support to be provided to carers in the long-term after stroke^89^.

### Stability of Psychological Outcomes in Long Term Stroke

Domain-general cognitive impairment on the OCS and MoCA were found to be statistically stable. However, when considering MCID change using an anchor-based estimate for the MoCA, 42% of participants declined and 20% improved. Discourse language, executive function and perceptuomotor abilities were statistically most variable across timepoints. The discourse language task was based on visual stimuli, and practice effects^90^ are likely to have contributed to variability. Similarly, executive function measured by the Hayling test improved over one year. However, only response initiation time decreased, suggesting participants improved in response speed only. This may partially explain why 60% demonstrated MCID improvements. Attention and memory abilities were statistically stable, consistent with previous findings^91, 92^. However, 20% - 49% demonstrated decline on memory tasks using MCID metrics. Exploring whether MCID changes in either direction are genuine or simply measurement error requires further research. Although we observed mean shifts in scores, impairment status was found to be stable over time. This could indicate that while improvement can occur in the long-term, individuals may not reach a status of “recovery.”

Self- and carer-reported depression and anxiety showed no statistically significant change over time. Emotional outcomes >2 years post-stroke may therefore be particularly stable. Participants may report higher distress in early stroke regardless of risk for chronic distress. Reviews note declines in depression and anxiety cases in the first year post-stroke^32^, however beyond one-year estimates remain stable^9^. Apathy and sleep levels also did not statistically change, aligning with previous work^32, 93^. Similarly, across these measures 50% - 77% showed no MCID change. Thus, much like emotional outcomes, apathy and sleep are long-term targets for intervention. Though stroke-related quality of life (98% no MCID change) and functional abilities (54% - 61%) were highly consistent between assessment timepoints, there were improvements on the EQ-5D-5L (37% MCID improvement), suggesting that regardless of persistent symptoms, some individuals may experience improvements in the very long-term.^94^

The only self-reported outcome to decline over the period of one year was fatigue. Investigating causes of worsening fatigue is a top unmet need reported by both stroke survivors and clinicians^16^. While fatigue levels are not thought to be affected by time post-stroke^86^, these data suggest there may be an eventual worsening of fatigue in the very long-term. Whilst replication is warranted, exploring factors relating to fatigue and intervention development is necessary. Likely, there are differing prevalence rates of fatigue subtypes (e.g., physical, emotional, and mental). Establishing the degree to which different subtypes of fatigue impact daily function, and how each subtype relates to outcomes, would be an asset in long-term fatigue management post-stroke.

## Clinical Implications

### Frequency vs Impact on Quality of Life

Whilst services should anticipate which psychological outcomes are most likely to need clinical attention, adequate time and effort should also go towards supporting those with less prevalent outcomes that may also affect quality of life. For example, though sleep difficulties were one of the least prevalent outcomes here, this does not presume that it has no impact on day-to-day functioning. Similarly, although clinically significant fatigue rates were double that of depression, depression more strongly correlated with stroke-related quality of life. Further, the ways in which quality of life is affected by psychological outcomes is important to understand – greater cognitive impairment was only correlated with stroke-specific quality of life rather than general quality of life, indicating there may be aspects of quality of life that may not be strongly impacted by cognition.

### Right Treatments at the Right Time

Findings suggest that the majority of long-term outcomes will remain stable relative to early stroke^33^. However, some stroke survivors demonstrated improvement, contradicting the notion that improvements only occur in the first-year post-stroke. This is further supported by the recent findings of long-term improvement with physical interventions in chronic stroke^95^. This sends a strong positive message that conducting interventions within chronic stroke may be as valuable as interventions in early stroke. Further, we found evidence of significant worsening of fatigue indicating that interventions in chronic stroke may also be valuable to prevent longer-term decline.

### Impact of Participant Attrition

Neither demographic variables, nor nature or severity of cognitive impairment differed between those lost to attrition and those retained. In combination with reasons for attrition (death, poor health, too busy to take part), attrition was likely not due to study-related factors making participation for stroke survivors difficult. However, as individuals lost to attrition self-reported overall poorer SF-SIS functioning, worse ADLs, and greater emotional distress, prevalence in these measures may be less representative.

## Limitations

Due to the COVID-19 pandemic, all assessments in OX-CHRONIC were conducted remotely. Though remote administration of the OCS (Tele-OCS) has been validated^96^, this format did not allow for apraxia impairments assessment. Though evidence suggests remote assessment of neuropsychological tests are comparable to in-person^97^, time-based metrics may be especially more variable via remote assessment. While time post-stroke did not correlate with key outcomes, OX-CHRONIC comprised a wide range of participants from 2 to 9 years post-acute event. As data collection was conducted during the pandemic, it is possible that this experience affected performance on study measures in unknown ways.

## Conclusions

Cognitive impairment was present in 45% of chronic stroke survivors. Domain-specific impairments in attention and executive function were the most common in this chronic sample. Memory impairments were the most stable, while discourse language abilities were more variable. There were high rates of depression, anxiety, fatigue, and apathy, and these outcomes correlated with worse quality of life in long-term stroke. This study elucidates the frequency of an array of psychological outcomes in chronic stroke survivors. These findings highlight that psychological consequences of stroke are prevalent and warrant attention in community-based stroke care.

## Data Availability

All data produced in the present study are available online at osf.io/y2mev

## Declarations

### Ethical Approval and Consent to Participate

This study received ethical approval from the Health Research Authority - South Central Berkshire Research Ethics Committee approved this study (REC Reference: 19/SC/0520)

### Consent for Publication

Participants provided informed consent for their data to be used for publication purposes.

### Availability of Data and Materials

The dataset supporting the conclusions of this article is available in the study-specific Open Science Framework repository: osf.io/y2mev

### Competing Interests

ND is a developer of the Oxford Cognitive Screen but does not receive any remuneration from its use. TJQ chairs the DMC for a vascular cognitive impairment trial supported by NovoNordisk; TJQ has provided outcomes assessment and advisory board input for trials in cognition for Novartis, NovoNordisk.

### Funding

This study was funded by a Priority Programme Grant from the Stroke Association (SA PPA 18/100032). ND (Advanced Fellowship NIHR302224) is funded by the National Institute for Health Research (NIHR). The project was supported by the National Institute for Health Research (NIHR) Oxford Health BRC. The views expressed in this publication are those of the author(s) and not necessarily those of the NIHR, NHS or the UK Department of Health and Social Care.

### Authors Contributions

ND, EM, TJQ, STP, A Kuppuswamy, ST, HD conceptualised the study and contributed to securing funding. ND, OAW, EM, EGC, HD, STP, A Kuppswamy, and TJQ contributed to protocol development. ND, OAW, and EM contributed to gaining ethical approval. ND, OAW, A Kusec and EGC contribute to study methodology. AK, CC, PW, EGC, EM, BD, and AD contributed to patient recruitment and data curation. ND, A Kusec and OAW conducted project administration. ND, TJQ, STP, A Kuppuswammy, ST, HD, TJ, and BA provided study supervision and management. A Kusec conducted all analyses and wrote the first manuscript draft. All authors reviewed and edited the manuscript and approved the final version of the manuscript.

## Acknowledgements

The authors would like to thank Audrey Bowen (chair), Avril Drummond, Richard Francis, Anna Volkmer, and Jeremy Dearling for providing study oversight as part of the OX-CHRONIC steering committee.

## List of Abberviations

AES: Apathy Evaluation Scale
CSI: Caregiver Strain Index
HADS: Hospital Anxiety and Depression Scale
ICECAP: ICEpop Capability Measure for Adults
IQ-CODE: Informant Questionnaire on Cognitive Decline in the Elderly
EQ-5D-5L: EuroQol 5-Dimesions 5-Levels
FDR: False Discovery Rate
FSS: Fatigue Severity Scale
GDS: Geriatric Depression Scale
MCID: Minimal Clinically Important Difference
MoCA: Montreal Cognitive Assessment
MMSE: Mini-Mental State Examination
NIHSS: National Institutes of Health Stroke Scale
OCS: Oxford Cognitive Screen
SCI: Sleep Condition Indicator
SF-SIS: Stroke Impact Scale Short Form
WHOQOL-BREF: World Health Organization Quality of Life Brief Version

## Footnotes

1 Activity monitor data will be reported elsewhere.

## Notes

### Clinical Protocols

https://journals.sagepub.com/doi/full/10.1177/23969873211046120

### Author Declarations

The Health Research Authority - South Central Berkshire Research Ethics Committee approved this study (REC Reference: 19/SC/0520)

### Summary of Updates

Corrected typos, updated abstracted slightly

