## Supplementary Materials for "Long-term psychological outcomes following stroke: The OX-CHRONIC study"

**Study Measures and Validated Cut Scores**

| **OX-CHRONIC Neuropsychological Assessment Measures** | | | |
| --- | --- | --- | --- |
| **Focus** | **Measure** | **Cut Score Metric** | **Cut Score** |
| **Domain-General Cognition** | OCS Subtask Total  Montreal Cognitive Assessment | Total Subtasks Impaired  Total Score | --  <26 & <22 |
| **Language** | OCS Picture Naming  OCS Semantics  OCS Sentence Reading  Cookie Theft Task  Boston Naming Test  DKEFS Letter Fluency  DKEFS Category Fluency | Total Correct  Total Correct  Total Correct  Complexity Ratio  Total Correct Without Cue  Total Correct Words  Total Correct Words | <3  <3  <14  <0.31  <11  <11 – <18  <14 – <24 |
| **Executive Function** | OCS Mixed Trails  DKEFS Trail Making Test A  DKEFS Trail Making Test B  Hayling Sentence Completion Test  OCS-Plus Trails | Total Accuracy  Total Time (seconds)  Total Errors  Overall Scaled Score  Executive Score | <7  >273  >3  <2  <40.54 |
| **Memory** | OCS Orientation  OCS Recall & Recognition  OCS Episodic Recognition  Digit Span Forwards  Digit Span Backwards  WMS-III Logical Memory Test I  WMS-II Logical Memory Test II  Picture Memory Test | Total Correct  Total Correct  Total Correct  Total Correct Recalls  Total Correct Recalls  Total Correct (Story A)  Total Correct (Story A)  Total Objects Correct | <4  <3  <3  <6 – <4  <4 – <3  <2 – <6  <3  <9 |
| **Visuospatial Attention** | OCS Broken Hearts  BIT Star Cancellation Task | Total Accuracy  Total Correct | <42  <51 |
| **Number Processing** | OCS Number Writing  OCS Calculation | Total Score  Total Correct | <3  <3 |
| **Perceptuomotor Abilities** | OCS-Plus Figure Copy  OCS-Plus Figure Recall  Rey-Osterrieth Complex Figure Copy  Rey-Osterrieth Complex Figure Recall | Total Score  Total Score  Total Score  Total Score | <44.41  <26.22  <13.5– 22.5  <3.5 – 4.5 |
| **OX-CHRONIC Self-Report Questionnaire Measures** | | | |
| **Participant Rated** | | | |
| **Domain** | **Measure** | **Metric Reported** | **Cut off** |
| **Post-Stroke Abilities** | Barthel – Short Form  Modified Rankin Scale  Nottingham Extended ADL Scale | Total Score  Scale score  Total Score | --  ≥3  -- |
| **Subjective Cognition** | Cognitive Failures Questionnaire  Cognitive Reserve Index | Total Score  Standardized Total | ≥43  -- |
| **Emotional Distress** | HADS-Depression  HADS-Anxiety  Geriatric Depression Scale | Subscale Total  Subscale Total  Scale Total | ≥8  ≥8  ≥5 |
| **Extended Outcomes** | Fatigue Severity Scale  Apathy Evaluation Scale  Sleep Condition Indicator | Scale Total  Scale Total  Scale Total | ≥36  ≥34  ≤16 |
| **Quality of Life** | Stroke Impact Scale-Short Form  EuroQol-5 Dimensions-5 Levels  WHO Quality of Life Abbreviated  ICEpop Capability Measure Adults | Scaled Total Score  Health Rating Score  Total Score  Total Score | --  --  --  -- |
| **Carer Rated** | | | |
| **Cognitive Decline** | Informant Questionnaire on Cognitive Decline in the Elderly | Averaged Total Score | ≥3.27 |
| **Emotional Distress** | Informant Assessed Geriatric Depression Scale | Total Score | ≥5 |
| **Carer Strain** | Modified Caregiver Strain Index | Total Score | ≥7 |

**Supplementary Table 1.** Neuropsychological assessments, self-report, and carer measures used in OX-CHRONIC with test metrics and cut off scores used to determine cognitive impairment. Where a range of impairment cutoffs are listed, this is due to some neuropsychological assessments have age-specific cutoffs for cognitive impairment.

OCS: Oxford Cognitive Screen; DKEFS: Delis-Kaplan Executive Function Systems; HSCT: Hayling Sentence Completion Test; WMS: Wechsler Memory Scale; BIT: Behavioural Inattention Test; ADL = Activities of Daily Living; HADS: Hospital Anxiety and Depression Scale

**Attrition Analysis**

A total of 15 participants were lost to attrition from Wave 1 to Wave 2 of OX-CHRONIC (*Supplementary Table 2*). Study attrition was not related to participant age at Wave 1 (*t* = 0.34, *p* = 0.73), sex (ꭓ^2^ = 0.42*, p* = 0.52), handedness (ꭓ^2^ = 0.37*, p* = 0.83), or whether individuals had an ischaemic or haemorrhagic stroke (ꭓ^2^ = 0.11*, p* = 0.75). Additionally, participants lost to attrition and those retained had a similar number of years of education (*t* = 0.58, *p* = 0.56), similar NIHSS stroke severity scores (*t* = -0.02, *p* = 0.99), and were similar number of years post-stroke (*t* = -0.62, *p* = 0.54).

In terms of cognitive scores, participants lost to attrition did not have a greater number of tasks impaired on the Oxford Cognitive Screen compared to those retained (Welch’s *t* = -0.71, *p* = 0.49). Between those retained and those lost to attrition, there were approximately equivalent proportions of individuals with language impairments (11.49% vs 9.10%; ꭓ^2^ = 0.06*, p* = 0.81), memory impairments (8.04% vs 9.10%; ꭓ^2^ = 0.01*, p* = 0.91), attention impairments (25.29% vs 27.27%; ꭓ^2^ = 0.02*, p* = 0.89), and number processing impairments (14.11% vs 9.10%; ꭓ^2^ = 0.21*, p* = 0.65). There appeared to be a greater proportion of participants lost to attrition with executive function impairments compared to those retained (27.27% vs 10.34% participants respectively), but this was not statistically significant (ꭓ^2^ = 2.60*, p* = 0.11).

In terms of psychological scores, participants lost to attrition were more likely to have lower scores on the SF-SIS at Wave 1 compared to those retained, suggesting poorer overall functioning (*t* = 2.29, *p* = 0.02). In specific SF-SIS domains, participants lost to attrition were more likely to have lower levels of ADLs (*t* = 2.31, *p* = 0.02) and poor emotional wellbeing (*t* = 2.09, *p* = 0.04). All other SF-SIS domains appeared lower in those lost to attrition, though these did not cross a statistical significance threshold (*t*s = 0.56 – 1.85, *p*s = 0.07 – 0.58).

Results remained the same when comparing participants who withdrew due to ill health and declining participation only (i.e., excluding attrition due to death) in terms of demographic (*p*s *=* 0.45 – 1), cognitive factors (*p*s = 0.19 – 1), or self-reported SF-SIS scores (*p*s = 0.06 – 0.82).

| **Measure** | **Retained (*N* = 90)** | **Withdrawn (*N* = 15)** | **Test Statistic** |
| --- | --- | --- | --- |
| **Demographics**  Age  Sex  Handedness  Stroke Type  Years of Education  Acute NIHSS Score  Time Post-Stroke | *M =* 73.10 (12.87)  57.78% Male  90.00% Righthanded  83.33% Ischaemic  *M =* 14.15 (3.97)  *M =* 7.38 (6.35)  *M =* 4.52 (2.09) | *M =* 71.84 (14.21)  66.67% Male  93.33% Righthanded  86.67% Ischaemic  *M =* 13.50 (4.31)  *M =* 7.42 (5.84)  *M =* 4.89 (2.34)) | *t =* 0.34  ꭓ^2^ *=* 0.42  ꭓ^2^ *=* 0.37  ꭓ^2^ *=* 0.11  *t =* 0.58  *t =* -0.02  *t =* -0.62 |
| **Cognitive Functioning**  OCS Number of Tasks Impaired  Language Impairment  Memory Impairment  Attention Impairment  Number Processing Impairment  Executive Function Impairment | *M =* 0.84 (1.22)  11.49% Impaired  8.04% Impaired  25.29% Impaired  14.11% Impaired  10.34% Impaired | *M =* 1.36 (2.42)  9.10% Impaired  9.10% Impaired  27.27% Impaired  9.10% Impaired  27.27% Impaired | Welch’s *t =* -0.71  ꭓ^2^ *=* 0.06  ꭓ^2^ *=* 0.01  ꭓ^2^ *=* 0.02  ꭓ^2^ *=* 0.21  ꭓ^2^ *=* 2.60 |
| **Stroke-Related Functioning**  SIS Total Scaled Score  Arm Functioning  Hand Functioning  Mobility  Communication  ADLs  Memory  Emotions  Participation | ***M =* 74.52 (18.92)**  *M =* 3.77 (1.05)  *M =* 3.89 (1.28)  *M =* 3.89 (1.17)  *M =* 4.43 (0.75)  ***M =* 4.02 (1.24)**  *M =* 4.01 (0.91)  ***M =* 4.28 (0.99)**  *M =* 3.55 (1.39) | ***M =* 61.88 (23.95)**  *M =* 3.27 (1.10)  *M =* 3.40 (1.50)  *M =* 3.27 (1.39)  *M =* 4.13 (1.13)  ***M =* 3.20 (1.47)**  *M =* 3.87 (1.06)  ***M =* 3.67 (1.35)**  *M =* 3.00 (1.41) | ***t =* 2.29***  *t =* 1.69  *t =* 1.33  *t =* 1.85  *t =* 0.99  ***t =* 2.31***  *t =* 0.56  ***t =* 2.09***  *t =* 1.42 |

**Supplementary Table 2.** Mean and frequency statistics of demographic, cognitive, and stroke outcome data for participants that were retained across time points (*N* = 90) and those who withdrew (*N* = 15).

OCS = Oxford Cognitive Screen; SIS = Stroke Impact Scale; ADLs = Activities of Daily Living

* *p* < 0.05

|  |  | **WAVE 1 (*N* = 98)** | | | **WAVE 2 (*N =* 85)** | | |
| --- | --- | --- | --- | --- | --- | --- | --- |
| **Domain** | **Measure** | **Median** | **Min - Max** | **Impairment Status – N (% [95% CI])** | **Median** | **Min – Max** | **Impairment Status – N (% [95% CI])** |
| **LANGUAGE**  Picture Naming  Semantics  Sentence Reading | Accuracy  Accuracy  Accuracy | 4  3  15 | 1 – 4  3 – 3  1 – 15 | 4 (4.1% [0.2 – 7.9])  0 (0.00% [0 - 0])  7 (7.1% [2.0 - 12.2]) | 4  3  15 | 2 – 4  2 – 3  8 – 15 | 3 (3.5% [-0.4 – 7.5])  1 (1.2% [-1.2 – 3.5])  9 (10.6% [4.1 – 17.1]) |
| **EXECUTIVE FUNCTION**  Circles Trails  Squares Trails  Mixed Trails | Accuracy  Time (seconds)  Accuracy  Time (seconds)  Accuracy  Time (seconds)  Executive Score | 6  12  6  13  13  39  -1 | 0 – 6  3 – 85  0 – 6  2 – 73  0 – 13  14 – 169  -7 – 11 | --  --  --  --  14 (14.3% [7.4 – 21.2])  --  12 (12.2% [5.8 – 18.7]) | 6  12  6  12.50  13  38  -1 | 0 – 6  5 – 102  4 – 6  4 – 50  4 – 13  10 – 157  -2 – 8 | --  --  --  --  4 (4.7% [0.2 – 9.2])  --  4 (4.7% [0.2 – 9.2]) |
| **MEMORY**  Orientation  Verbal Recall & Recognition    Episodic Recognition | Accuracy  Free Recall  Recall + Recognition  Accuracy | 4  3  4  4 | 1 – 4  0 – 4  0 – 4  2 – 4 | 4 (4.1% [0.2 – 7.9])  --  5 (5.2% [0.8 – 9.5])  1 (1.0% [-0.9 – 3.0]) | 4  3  4  4 | 2 – 4  0 – 4  1 – 4  3 – 4 | 5 (5.9% [0.9 – 10.9])  --  3 (3.5% [-0.4 – 7.5])  0 (0.00% [0 – 0]) |
| **ATTENTION**  Broken Hearts | Accuracy  Time (seconds)  Left Gap Total  Right Gap Total  Space Asymmetry  Egocentric Neglect  Space Neglect Left  Space Neglect Right  Object Asymmetry  Allocentric Neglect  Object Neglect Left  Object Neglect Right | 46  122  0  0  0  --  --  --  0  --  --  -- | 9 – 50  34 – 180  0 – 14  0 – 7  -13 – 9  --  --  --  -3 – 14  --  --  -- | 21 (21.4% [13.3 – 29.6])  --  --  --  --  8 (8.2% [2.74 – 13.6])  9 (9.3% [3.54 – 15.0])  5 (5.2% [0.77 – 9.5])  --  8 (8.2% [2.7 – 13.6])  4 (4.1% [0.2 – 7.9])  4 (4.1% [0.2 – 7.9]) | 46  124.50  0  0  0  --  --  --  0  --  --  -- | 15 – 50  53 – 190  0 – 11  0 – 10  -12 – 8  --  --  --  -3 – 9  --  --  -- | 23 (27.1% [17.6 – 36.5])  --  --  --  --  7 (8.2% [2.4 – 14.1])  3 (3.5% [-0.4 – 7.5])  8 (9.4% [3.2 – 15.6])  --  7 (8.2% [2.4 – 14.1])  3 (3.5% [-0.4 – 7.5])  4 (4.7% [0.2 – 9.2]) |
| **NUMBER PROCESSING**  Number Writing  Calculation | Accuracy  Accuracy | 3  4 | 1 – 3  2 – 4 | 12 (12.2% [5.8 – 18.7])  3 (3.1% [-0.4 – 6.5]) | 3  4 | 0 – 3  2 – 4 | 14 (16.5% [8.6 – 24.4])  4 (4.7% [0.2 – 9.2]) |

**Supplementary Table 3.** Descriptive and impairment prevalence statistics per domain on the Oxford Cognitive Screen (OCS) at Wave 1 (*N* = 98) and Wave 2 (*N* = 85). Impairment scores were calculated using normative data published in Demeyere et al. (2015).

|  | | **WAVE 1 (*N* = 98)** | | | **WAVE 2 (*N* = 85)** | | |
| --- | --- | --- | --- | --- | --- | --- | --- |
|  | **Measure** | **Mean (SD)** | **Min – Max** | **Impairment Status**  **N (% [95% CI])** | **Mean (SD)** | **Min – Max** | **Impairment Status**  **N (% [95% CI])** |
| **DOMAIN-GENERAL COGNITION**  OCS  MoCA | Tasks Impaired  Total Score  ≤26  ≤22 | 0.90 (1.40)  23.56 (4.16) | 0 – 8  13 – 30 | 45 (45.9% [36.1 – 55.8])  64 (65.3% [55.9 – 74.7])  30 (30.6% [21.5 – 39.7]) | 0.99 (1.33)  22.98 (4.51) | 0 – 5  10 – 30 | 40 (47.1% [36.5 – 57.7])  57 (67.1% [57.1 – 77.1])  29 (34.1% [24.0 – 44.2]) |
| **LANGUAGE**  Cookie Theft Task  BNT  Letter Fluency  Category Fluency | Total Utterances  Empty Utterances (%)  Subclausal Utterances (%)  Single Clause Utterances (%)  Multiclause Utterances (%)  Agrammatic Deletions (%)  Complexity Index  Total Correct No Cue  Total Score  Set Loss Errors  Repetition Errors  Total Score  Set Loss Errors  Repetition Errors | 17.90 (10.25)  12.90 (11.01)  23.87 (15.23)  67.42 (14.58)  5.96 (8.16)  4.42 (7.29)  0.81 (0.23)  13.89 (1.79)  32.53 (14.94)  1.35 (1.66)  0.66 (1.18)  31.64 (10.00)  0.86 (1.35)  0.16 (0.78) | 4 – 74  0 – 55.6  0 – 64  24 – 100  0 – 27.77  0 – 37.5  0.17 –1.31  6 – 15  9 – 74  0 – 7  0 – 7  5 – 54  0 – 7  0 – 7 | --  --  --  --  --  --  1 (1.0% [-0.9 – 3.1])  4 (4.1% [0.2 – 7.9])  14 (14.3% [7.4 – 21.2])  --  --  8 (8.2% [2.7 – 13.6])  --  -- | 24.18 (11.79)  11.25 (8.85)  7.26 (7.05)  60.28 (12.88)  22.57 (12.67)  4.85 (6.52)  1.22 (0.24)  --  32.69 (14.88)  1.38 (1.54)  0.92 (1.11)  32.51 (11.09)  0.91 (1.39)  0.18 (0.62) | 6 – 68  0 – 43.86  0 – 28.57  28.57–87.50  0 – 58.62  0 – 33.33  0.12 – 1.96  **--**  6 – 78  0 – 5  0 – 7  11 – 57  0 – 4  0 – 7 | --  --  --  --  --  --  1 (1.2% [-1.1 – 3.5])  **--**  9 (10.7% [4.1 – 17.3])  --  --  7 (8.3% [2.5 – 14.2])  --  -- |
| **EXECUTIVE FUNCTION**  Trails A  Trails B    Hayling Test  OCS-Plus Mixed Trails | Total Accuracy  Time (seconds)  Sequencing Errors  Total Accuracy  Time (seconds)  Total Errors  Sequencing Errors  Set Loss Errors  Total Score  Part A – Time (seconds)  Part B – Time (seconds)  Converted Error Score  Circles Accuracy  Circles Time (seconds)  Squares Accuracy  Squares Time (seconds)  Mixed Accuracy  Mixed Time (seconds) | 23.81 (0.63)  49.91 (31.82)  0.25 (0.69)  18.54 (6.00)  122.42 (89.11)  2.95 (4.45)  1.60 (2.94)  1.35 (2.25)  12.03 (3.65)  36.82 (29.04)  85.12 (47.97)  12.08 (11.35)  6.45 (1.70)  16.82 (10.57)  6.69 (0.99)  17.72 (11.47)  10.55 (4.16)  68.90 (56.89) | 20 – 24  18 – 215  0 – 4  3 – 23  24 – 651  0 – 18  0 – 14  0 – 11  4 – 19  4 – 201  21 – 232  0 – 52  0 – 7  2 – 60  0 – 7  4 – 60  1 – 14  18 – 414 | --  11 (11.3% [5.1 – 17.6])  --  --  5 (5.2% [0.8 –9.5])  28 (28.9% [19.9 – 37.8])  --  --  30 (30.6% [21.5 – 39.7])  10 (10.2% [4.21 – 16.1])  17 (17.4% [9.85 – 24.9])  8 (8.2% [2.7 – 13.6])  --  --  --  --  15 (15.5% [8.3 – 22.6])  -- | 23.48 (1.89)  51.16 (31.65)  0.33 (0.73)  18.04 (5.55)  129.6(100.87)  2.17 (3.03)  1.98 (3.21)  4.14 (5.11)  13.60 (3.93)  24.29 (15.31)  68.62 (52.56)  11.43 (10.90)  6.81 (0.91)  15.87 (11.10)  6.75 (0.66)  15.09 (8.55)  10.61 (3.76)  60.58 (34.23) | 8 – 24  18 – 185  0 – 4  1 – 23  32 – 697  0 – 20  0 – 14  0 – 20  6 – 19  6 – 93  9 – 287  0 – 50  0 – 7  5 – 74  4 – 7  4 – 45  1 – 14  14 - 208 | --  8 (9.4% [3.2 – 15.6])  --  --  4 (4.7% [0.2 – 9.2])  34 (40.0% [29.6 – 50.4])  --  --  20 (23.5% [14.5 – 32.6])  2 (2.4% [-0.9 – 5.6])  10 (11.8% [4.9 – 18.6])  6 (7.1% [1.6 – 12.5])  --  --  --  --  23 (27.1% [17.6 – 36.5])  -- |
| **MEMORY**  Digit Span Forward  Digit Span Backward  Logical Memory – 1  Logical Memory – 2    Picture Memory | Total Score  Max Length  Total Score  Max Length  Total Score  Total Score  Total Score  Related Errors  Unrelated Errors | 7.49 (2.33)  6.18 (1.22)  5.90 (2.14)  4.49 (1.21)  12.37 (4.46)  10.67 (5.19)  9.93 (2.73)  0.47 (0.92)  0.02 (0.14) | 0 – 12  2 – 8  1 – 12  2 – 7  1 – 22  0 – 20  2 – 15  0 – 4  0 – 1 | 9 (9.2% [3.5 – 15.0])  --  4 (4.1% [0.2 – 7.9])  --  3 (3.1% [-0.3 – 6.5])  9 (9.3% [3.5 – 15.0])  26 (26.5% [17.8 – 35.3])  --  -- | 7.25 (2.57)  6.07 (1.37)  5.89 (2.36)  4.33 (1.31)  12.24 (4.50)  10.73 (4.96)  --  **--**  **--** | 0 – 12  2 – 8  0 – 12  1 – 7  2 – 21  0 – 21  **--**  **--**  **--** | 7 (8.2% [2.4 – 14.1])  --  5 (5.9% [0.88 – 10.9])  --  2 (2.4% [-0.9 – 5.6])  6 (7.1% [1.6 – 12.5])  --  --  -- |
| **VISUOSPATIAL ATTENTION**  Star Cancellation | Total Score  Time (seconds)  Total Left  Total Right  Laterality Index  Left Spatial Neglect  Right Spatial Neglect | 52.19 (5.70)  98.24 (48.53)  26.03 (3.05)  26.15 (2.96)  0 (0.09)  --  -- | 8 – 54  38 – 302  7 – 27  1 – 27  -0.75-0.28  --  -- | 18 (18.6% [10.9 – 26.3])  --  --  --  --  5 (5.2% [0.8 – 9.5])  0 (0.0% [0 – 0]) | 52.76 (2.26)  98.51 (43.46)  26.31 (1.67)  26.45 (1.20)  0 (0.04)  --  -- | 41 – 54  40 – 228  14 – 27  20 – 27  -0.08-0.32  --  -- | 14 (16.5% [8.6 – 24.4])  --  --  --  --  2 (2.4% [-0.9 – 5.6])  0 (0.00% [0 – 0]) |
| **PERCEPTUOMOTOR ABILITIES**  OCS-Plus Figure  ROCF | Copy – Total Score  Copy – Time (seconds)  Recall – Total Score  Recall – Time (seconds)  Copy – Total Score  Copy – Time (seconds)  Recall – Total Score  Recall – Time (seconds) | 54.99 (5.85)  60.92 (39.13)  41.46 (10.57)  58.98 (39.67)  26.68 (6.38)  172.26 (103.88)  12.79 (7.35)  102.43 (70.43) | 29 – 60  16 – 244  16 – 59  3 – 308  4 – 36  61 – 828  0 – 29  12 – 460 | 6 (6.1% [1.4 – 10.9])  --  6 (6.1% [1.4 – 10.9])  --  10 (10.2% [4.2 – 16.2])  **--**  8 (8.3% [2.9 – 13.8])  **--** | 54.07 (5.19)  56.39 (28.24)  39.81 (9.65)  63.56 (34.63)  24.04 (6.35)  165.2(107.34)  12.42 (6.61)  112.7(71.79) | 35 – 60  21 – 142  18 – 57  21 – 216  5 – 34  51 – 892  1.5 – 28  21 – 385 | 5 (5.9% [0.9 – 10.9])  --  9 (10.6% [4.1 – 17.1])  --  17 (20.2% [11.7 – 28.8])  --  4 (9.4% [3.2 – 15.6])  -- |

**Supplementary Table 4.** Descriptive and impairment prevalence statistics per domain using in-depth neuropsychological assessments at Wave 1 (*N* = 98) and Wave 2 (*N* = 85). Impairment scores used are shown in *Supplementary Table 1.*

OCS = Oxford Cognitive Screen; MoCA = Montreal Cognitive Assessment; BNT = Boston Naming Test; ROCF = Rey-Osterrieth Complex Figure

|  | **Wave 1 Proportion Impaired** | **Wave 2 Proportion Impaired** | **FDR-adjusted χ^2^** |
| --- | --- | --- | --- |
| **Domain-General Cognition**  OCS Tasks Impaired  MoCA (<26)  MoCA (<22) | 0.4592  0.6531  0.3061 | 0.4706  0.6706  0.3412 | 0.02 (*p* = 0.97)  0.06 (*p* = 0.89)  0.26 (*p* = 0.96) |
| **Language**  Cookie Theft Complexity  Letter Fluency Total  Category Fluency Total | 0.0103  0.1429  0.0816 | 0.0120  0.1071  0.0833 | 0.01 (*p =* 0.97)  0.06 (*p* = 0.89)  0.26 (*p* = 0.96) |
| **Executive Function**  Trails A Accuracy  Trails B Accuracy  Hayling Test Total  OCS-Plus Mixed Trails | 0.1134  0.2887  0.3061  0.1546 | 0.0941  0.4000  0.2353  0.2706 | 0.52 (*p* = 0.96)  0.01 (*p* = 0.96)  1.13 (*p* = 0.89)  3.18 (*p* = 0.66) |
| **Memory**  Digit Span Forwards  Digit Span Backwards  Logical Memory I  Logical Memory II | 0.0928  0.0408  0.0310  0.0928 | 0.0825  0.0588  0.0238  0.0706 | 0.05 (*p* = 0.97)  0.32 (*p* = 0.96)  0.08 (*p* = 0.97)  0.27 (*p* = 0.97) |
| **Visuospatial Attention**  Star Cancellation Total | 0.1856 | 0.1647 | 0.11 (*p* = 0.97) |
| **Perceptuomotor Abilities**  OCS-Plus Figure Copy  OCS-Plus Figure Recall  ROCF Copy  ROCF Recall | 0.0612  0.0612  0.1020  0.0833 | 0.0588  0.1059  0.2024  0.0941 | 0.01 (*p =* 0.97) 1.20 (*p =* 0.89)   3.45 (*p* = 0.66)  0.92 (*p* = 0.89) |

**Supplementary Table 5.** Chi-square evaluations of whether proportion of individuals impaired differs across time points. False discovery rate adjustments on p-values from chi-square tests.

|  | **Measure** | ***N*** | **Mean (SD)** | **Min - Max** | **Clinically Significant –**  **N (% [96% CI])** |
| --- | --- | --- | --- | --- | --- |
| **POST-STROKE ABILTIES**  Modified Rankin Scale^1^  Barthel-Short Form^1^  Nottingham Extended ADL^1^ | Score  Total Score  Total Score | 100  99  100 | 1.79 (1.27)  6.93 (1.58)  47.77 (16.26) | 0 – 5  0 – 8  4 – 66 | 30 (28.6% [19.7 – 37.4])  --  -- |
| **SUBJECTIVE COGNITION**  Cognitive Failures Questionnaire^2^  Cognitive Reserve Index^2^ | Total Score  Distractibility  Forgetfulness  False Triggering  Total Score  Education  Work Activity  Leisure | 88  90  90  87  85  85  85  85 | 33.60 (17.93)  10.54 (5.78)  13.19 (6.80)  9.02 (5.91)  128.50 (19.51)  121.37 (14.58)  109.56 (22.99)  133.34 (21.78) | 1 – 89  0 – 27  1 – 29  0 – 26  86.68 – 191  83.83– 159.92  74 – 220  92 – 202.47 | 29 (32.2% [22.5 – 41.9])  --  --  --  --  --  --  -- |
| **EMOTIONAL DISTRESS**  Hospital Anxiety and Depression Scale^1^  Geriatric Depression Scale^1^ | Depression Total  Possible Depression  Probable Depression  Anxiety Total  Possible Anxiety  Probable Anxiety  Total Score  Mild Depression  Moderate Depression  Severe Depression | 98  98  101 | 4.97 (3.98)  5.23 (4.02)  4.22 (4.10) | 0 – 21  0 – 18  0 – 15 | 23 (23.5% [15.1 – 31.9])  11 (11.2% [4.9 – 17.5])  22 (22.5% [14.2 – 30.7])  12 (12.2% [5.8 – 18.7])  34 (33.7% [24.4 – 42.9])  16 (15.8% [8.7 – 22.9])  9 (9.9% [4.1 – 15.7]) |
| **EXTENDED OUTCOMES**  Apathy Evaluation Scale^1^  Fatigue Severity Scale^1^  Sleep Condition Indicator^1^ | Total Score  Total Score  Total Score | 101  101  100 | 32.36 (10.21)  35.56 (15.34)  23.29 (8.04) | 18 – 60  9 – 63  0 – 32 | 41 (40.6% [31.0 – 50.2])  52 (51.5% [41.7 – 61.2])  21 (21.0% [13.0 – 28.9]) |
| **QUALITY OF LIFE**  Stroke Impact Scale-Short Form^1^  Stroke Impact Scale-Long Form^2^  WHO-Quality of Life – Abbreviated^2^    ICECAP-A Capability Measure^1^  EQ5D-5L Quality of Life^1^ | Total Score (Scaled)  Arm Function (Scaled)  Memory (Scaled)  Emotion (Scaled)  Communication (Scaled)  ADL (Scaled)  Mobility (Scaled)  Hand Function (Scaled)  Participation (Scaled)  Stroke Recovery Score  Overall  Physical  Psychological  Social  Disability Module  Total Score  Stability  Attachment  Autonomy  Achievement  Enjoyment  Mobility  Self-Care  Usual Activities  Pain/Discomfort  Anxiety/Depression  Health Rating | 102  88  90  88  90  87  89  88  82  88  83  88  90  77  90  101  100  101  101  101  101  101  101  101  100  100  99 | 72.66 (20.12)  69.94 (24.06)  78.19 (20.64)  71.45 (17.87)  83.25 (18.33)  79.20 (20.31)  74.41 (23.00)  71.51 (31.64)  70.32 (25.92)  71.91 (21.63)  7.22 (1.65)  26.47 (5.44)  22.90 (3.92)  10.36 (2.59)  51.98 (8.55)  15.65 (2.90)  3.06 (0.89)  3.60 (0.60)  3.06 (0.85)  2.77 (0.84)  3.16 (0.73)  2.40 (1.22)  1.72 (0.99)  2.24 (1.17)  2.06 (0.83)  1.59 (0.81)  68.61 (18.83) | 9.38 – 100  0 – 100  14.29 – 100  8.33 – 100  17.86 – 100  27.50 – 100  11.11 – 100  0 – 100  6.25 – 100  20 – 100  3 – 10  11 – 34  7 – 30  3 – 15  25 – 65  8 – 20  1 – 4  2 – 4  1 – 4  1 – 4  1 – 4  1 – 5  1 – 5  1 – 5  1 – 4  1 – 5  10 – 100 | --  --  --  --  --  --  --  --  --  --  --  --  --  --  --  --  --  --  --  --  --  --  --  --  --  --  -- |
| **CARER MEASURES**  Caregiver Strain Index^1^  Informant Geriatric Depression Scale^1^  Informant Cognitive Decline^1^ | Total Score  Total Score  Total Score (Scaled) | 68  70  74 | 2.76 (3.12)  5.18 (4.30)  3.23 (0.63) | 0 – 11  0 – 15  1 – 4.81 | 9 (13.2% [5.2 – 21.3])  35 (50.0% [38.3 – 61.7])  29 (39.2% [28.1 – 50.3]) |

**Supplementary Table 6.** Descriptive statistics on self-reported and carer questionnaire measures. Clinical cutoff scores are in Supplementary Table 1.

WHO = World Health Organization; ADL = Activities of Daily Living; ICECAP-A = ICEpop Capability Measure for Adults; EQ5D-5L = EuroQol-5 Dimensions-5 Levels

^1^ Data presented was collected at Wave 1.

^2^ Data presented was collected at Wave 2.

| **Domain** | **Test Statistic^a^** | **Mean Difference (95% CI)** | **Cohen’s *d*** | **MCID**  **(0.5 SD)** | **Distribution MCID** | | | **Published MCID** | **Published MCID** | | |
| --- | --- | --- | --- | --- | --- | --- | --- | --- | --- | --- | --- |
|  |  |  |  |  | **Improve** | **Decline** | **No Change** |  | **Improve** | **Decline** | **No Change** |
| **Domain-General Cognition**  OCS Tasks Impaired  Montreal Cognitive Assessment | *V* = 403.50  *t* = 2.57 | -0.14 (-0.44, 0.16)  0.86 (0.29, 1.52) | -0.10  0.28 | 0.69  2.07 | 21.2%  11.7% | 24.7%  28.2% | 54.1%  60.1% | --  1.22^64^ | --  20.0% | --  42.3% | --  37.7% |
| **Language**  Cookie Theft Complexity  Letter Fluency Total  Category Fluency Total | ***t* = -12.13*****  *t* = 1.56  *t* = -0.41 | **-0.40 (-0.47, -0.34)**  1.04 (-0.28, 2.35)  -0.26 (-1.53, 1.01) | **-1.33**  0.17  -0.04 | 0.12  7.46  5.00 | **83.1%**  8.2%  16.4% | 3.6%  17.6%  15.3% | 13.3%  74.2%  68.3% | --  --  -- | --  --  -- | --  --  -- | --  --  -- |
| **Executive Function**  Trail Making Test A Accuracy  Trail Making Test B Accuracy  Hayling A Response Time  Hayling B Response Time  Hayling B Errors  Hayling Test Total  OCS-Plus Mixed Accuracy | *V* = 137  *V* = 992  ***t* = 4.24*****  *t* = 2.54  *t* = -0.35  ***t* = -4.45*****  *V* = 922.50 | 0.32 (-0.09, 0.74)  0.65 (-0.39, 1.71)  **13.38 (7.10, 19.66)**  13.27 (2.89, 23.65)  -0.36 (-2.43, 1.71)  **-1.42 (-2.06, -0.79)**  0.05 (-0.77, 0.87) | 0.17  0.14  **0.46**  0.28  -0.04  **-0.49**  0.01 | 0.32  2.99  14.52  23.98  5.67  0.92  2.08 | 7.1%  20.2%  **32.1%**  36.9%  24.1%  **60.2%**  17.8% | 15.4%  27.3%  4.7%  8.3%  28.9%  25.3%  21.4% | 77.5%  52.5%  63.2%  54.8%  47.0%  14.5%  60.8% | --  --  --  --  --  --  -- | --  --  --  --  --  --  -- | --  --  --  --  --  --  -- | --  --  --  --  --  --  -- |
| **Memory**  Digit Span Forwards  Digit Span Backwards  Logical Memory Immediate  Logical Memory Recall | *t* =1.11  *V* = 985.5  *t* = 0.83  *t* = 0.24 | 0.21 (-0.17, 0.59)  0.08 (-0.35, 0.51)  0.35 (-0.48, 1.17)  0.11 (-0.78, 0.99) | 0.12  0.04  0.09  0.03 | 1.16  1.07  2.23  2.59 | 3.5%  18.8%  23.5%  23.8% | 49.4%  20.0%  30.9%  29.7% | 47.1%  61.2%  45.6%  46.5% | --  --  --  -- | --  --  --  -- | --  --  --  -- | --  --  --  -- |
| **Visuospatial Attention**  Star Cancellation Task Total | *V* = 427.5 | -0.64 (-1.83, 0.55) | -0.12 | 2.85 | 9.5% | 4.7% | 85.8% | -- | -- | -- | -- |
| **Perceptuomotor Abilities**  OCS-Plus Figure Copy  OCS-Plus Figure Recall  ROCF – Copy  ROCF – Recall | *V* = 1834.50  *t* = 2.05  ***V* = 2581*****  *t* = 1.57 | 0.79 (-0.21, 1.78)  1.66 (0.05, 3.27)  **3.08 (2.11, 4.05)**  0.77 (-0.20, 1.73) | 0.17  0.22  **0.69**  0.17 | 2.92  5.28  3.19  3.67 | 12.9%  17.6%  3.5%  19.2% | 31.7%  31.7%  **40.4%**  22.8% | 55.4%  50.7%  56.1%  58.0% | --  --  --  -- | --  --  --  -- | --  --  --  -- | --  --  --  -- |
| **Post-Stroke Abilities**  Modified Rankin Scale Score  Barthel-3 item Total | *V* = 245.50  *V* = 217 | -0.22 (-0.43, -0.01)  -0.11 (-0.31, 0.09) | -0.23  -0.12 | 0.63  0.79 | 16.4%  24.7% | 29.4%  14.1% | 54.2%  61.2% | 1^65^  -- | 16.4%  -- | 29.4%  -- | 54.2%  -- |
| **Quality of Life**  SIS-Short Form Scaled Total  EQ5D-5L Health Rating | *t* = -0.48  ***V* = 936.50*** | -0.20 (-1.04, 0.64)  **-5.03 (-8.76, -1.31)** | -0.05  **-0.29** | 10.05  9.41 | 1.1%  **37.2%** | 1.1%  16.2% | 97.8%  46.6% | --  8.61^66^ | --  **37.2%** | --  16.2% | --  46.6% |
| **Emotional Distress**  HADS-Depression Total  HADS-Anxiety Total  GDS Total | *t* = -0.71  *t* = -1.15  *t* = 0.89 | -0.19 (-0.74, 0.35)  -0.37 (-1.02, 0.27)  0.21 (-0.26, 0.67) | -0.08  -0.13  0.09 | 1.99  2.01  2.04 | 24.7%  12.9%  16.1% | 25.8%  21.1%  6.9% | 49.5%  66.0%  77.1% | 2^67^  2^67^  2^68^ | 14.1%  12.9%  16.1% | 14.1%  21.1%  6.9% | 71.8%  66.0%  77.1% |
| **Extended Outcomes**  Apathy Evaluation Scale  Fatigue Severity Scale  Sleep Condition Indicator | *t* = 0.58  ***t* = -3.09***  *V* = 1468.00 | 0.31 (-0.75, 1.36)  **-3.90 (-6.41, -1.39)**  0.37 (-0.78, 1.54) | 0.06  **-0.33**  0.07 | 5.10  7.66  4.02 | 12.7%  15.1%  11.6% | 14.9%  **37.2%**  12.7% | 72.4%  47.7%  75.7% | --  4.05^69^  7.00^70^ | --  19.7%  4.6% | --  **45.3%**  5.8% | --  35.0%  89.6% |
| **Carer Measures**  Informant-GDS Total  IQ-CODE | *t* = -0.79  *V* = 936.50 | -0.21 (-0.79, 0.36)  0.15 (0.03, 0.27) | -0.10  0.34 | 2.15  0.31 | 9.1%  15.5% | 9.1%  5.1% | 81.8%  79.4% | 2^68^  -- | 9.1%  -- | 9.1%  -- | 81.8%  -- |

**Supplementary Table 7.** Complete case test statistics of neuropsychological assessment, self-report, and carer measures per domain between Wave 1 and Wave 2 (*N* = 90) including standardized mean difference scores and Cohen’s *d* effect size estimates. Statistical tests were alpha-adjusted using false discovery rate (FDR) corrections. Distribution-based MCIDs were estimated by calculating percentage of individuals whose difference in scores per measure between Wave 1 and Wave 2 were 0.5 standard deviations (SDs) above or below the mean of each measure at Wave 1. Where available, published anchor-based MCID estimates were used to compare to distribution-based MCIDs (referenced by number).

MCID: Minimal Clinically Important Difference; OCS: Oxford Cognitive Screen; ROCF: Rey-Osterrieth Complex Figure Copy; SIS: Stroke Impact Scale; EQ5D-5L: EuroQol-5 Dimensions-5 Levels; HADS: Hospital Anxiety and Depression Scale; GDS: Geriatric Depression Scale; IQ-CODE: Informant Questionnaire for Cognitive Decline in the Elderly

^a^For parametric data, paired t-tests were used. For non-parametric data, Wilcoxon tests were used.

* *p* < 0.05, ** *p* < 0.01, *** *p* < 0.001 following family-wise FDR corrections.

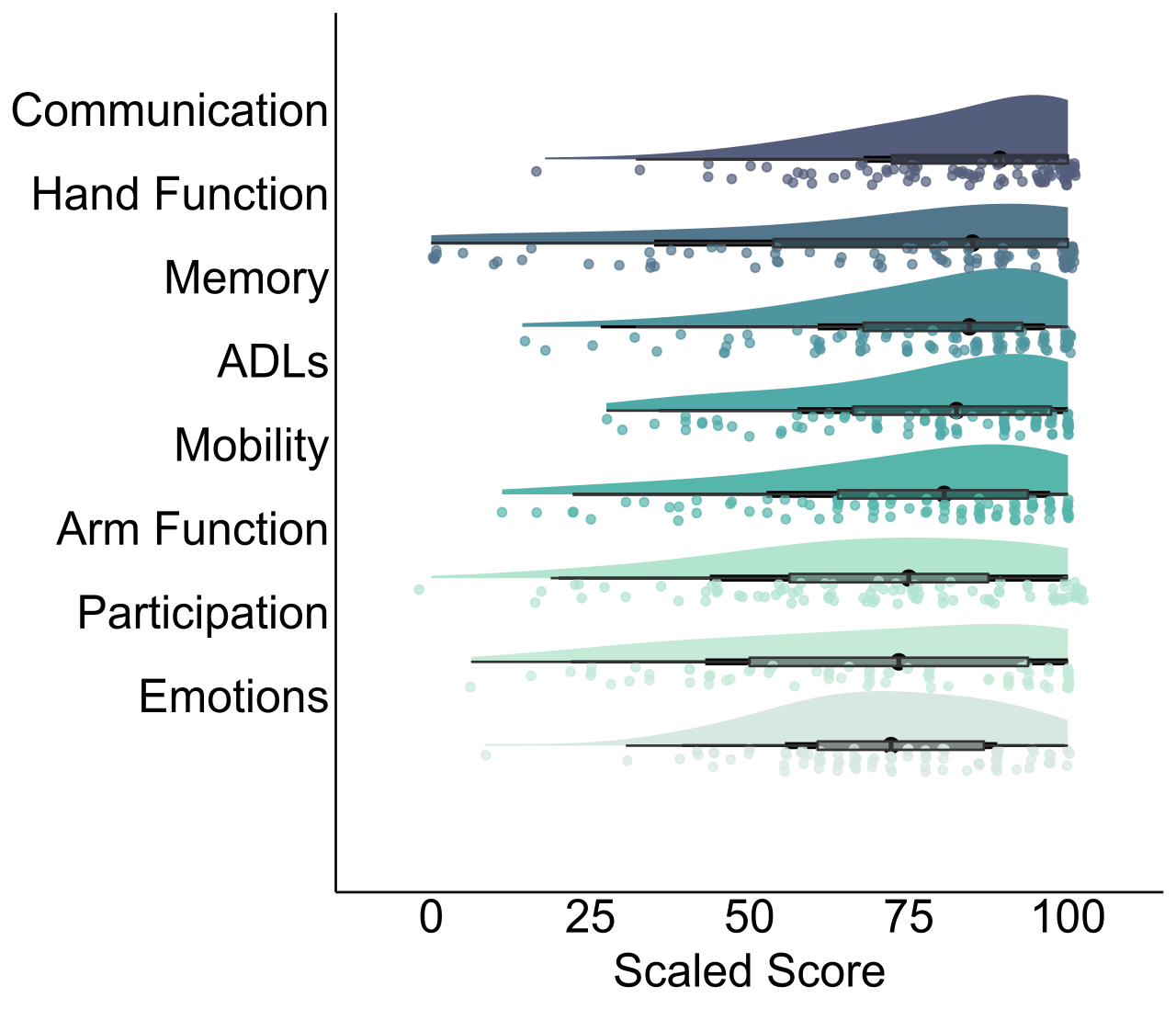

**Supplementary Figure 1.** Distributions of Stroke Impact Scale subscales in chronic stroke using complete cases at Wave 2 (*N* = 90). Median points with quantile ranges are shown within boxplots. Higher scaled scores represent better functioning. Scaled scores were highest in the domain of communication (median = 89.29), followed by hand function (median = 85), memory (median = 84.52), activities of daily living (median = 82.50), mobility (median = 80.56), arm function (median = 75), and participation (median = 73.44), with emotions having the lowest self-reported scores (median = 72.22).
